# Geospatial Accessibility to Primary and Community Health Centres in India

**DOI:** 10.1101/2025.05.13.25327502

**Authors:** Shreyas Patil, Aamir Miyajiwala, Anoushka Arora, Ankit Raj, Harsh Thakkar, Mukta S. Takalikar, Siddhesh Zadey

## Abstract

**Background:** Accessibility to public healthcare in India is poor. Data on access to public health facilities in terms of travel time and geographical scope are limited.

**Methods:** Primary health centre (PHC) and community health centre (CHC) locations were taken from the Geographic Information System dataset of the Pradhan Mantri Gram Sadak Yojana 2021. Motorized and walking travel-time friction surface rasters were taken from the Malaria Atlas Project 2020. We examined the densities, travel times, and Access Population Coverage (APC) for PHCs and CHCs in India up to the district level. For walking APCs, we used a 30-minute time threshold for PHCs and a 60-minute threshold for CHCs. For motorized APCs, we used a 60-minute threshold for PHCs and a 120-minute threshold for CHCs.

**Results:** In 2021, India had 23.22 PHCs and 3.07 CHCs per million people. Median PHC travel time was 79.53 (walking) and 16.77 minutes (motorised transport). CHC travel time was 227.53 (walking) and 36.23 minutes (motorised transport). PHC APCs for walking and motorised transport were 18.69% and 97.97%, respectively. CHC APCs for walking and motorised transport were 10.89% and 98.88%, respectively. Wide variations were seen in rural-urban analysis.

**Conclusion:** Our study found marked heterogeneity in timely geographic access, with access by walking being better in rural India and by motorised transport being better in urban India.

## 1 Background

India is currently the most populous country in the world, being home to over 1.4 billion individuals.(1) Safeguarding the health of this huge population has several challenges and requires immense resources with their commensurate distribution. Geographical factors like the size of the country, the 7th largest in the world, affect the distribution of these resources. Variations in terrain exacerbate these difficulties, particularly in hilly or mountainous regions.(2) The Alma-Ata declaration propounded the goal of “Health for All’’ recognising the importance of primary health care in attaining this objective.(3) In India, primary health care is rendered mainly by the primary health centres (PHCs), the first point of contact of the community with a trained medical officer. Above the PHCs in the continuum of care are the community health centres (CHCs), which are the first level of referral and provide specialist care. The Indian Public Health Standards (IPHS) 2022 recommend the availability of one PHC for every 50,000 people in urban areas, every 30,000 people in rural areas, and every 20,000 people in tribal/hilly areas. The recommendations for CHCs are that one facility must be available for every 80,000 people in tribal/hilly areas and for every 120,000 people in rural areas. For urban areas, the recommended population norms are one CHC for every 500,000 people in metro areas and every 250,000 people in non-metro regions.(4)

The number of health facilities in terms of a defined population size is referred to as the density of health facilities and is a measure of availability. On the other hand, accessibility is a spatial measure of ‘closeness’ or geographical proximity with an element of time.(5,6) A mere availability of health facilities will not fulfil the health needs of a population if geographic access is difficult due to reasons such as distance, terrain, or costs. Geographical distance and travel times are significant barriers to accessibility, universalization of health care, and thus utilisation of health services, particularly in rural settings.(6–13) Moreover, timely geographic access to healthcare is essential to ensure optimal health outcomes.(14–16) Longer travel times have a harmful influence on child mortality rates.(17,18) They are also associated with an increased risk of stillbirths, maternal morbidity, and mortality.(19–22) Increased distances to healthcare facilities can also negatively impact neonatal and maternal outcomes by decreasing the probability of women undergoing institutional delivery.(23,24) Distances do not necessarily indicate geographic access better due to various factors such as terrain. Time taken to travel distances, therefore, serves as a more accurate measure. Furthermore, longer travelling times impose additional burdens like higher travel costs and longer loss of wages.(25–27) Additionally, access, geography, and timeliness have been identified as forms of moral capital when healthcare systems are viewed as moral economies. Taking these into consideration can improve healthcare systems by making them more equitable, effective, and efficient.(28)

It is estimated that 44.2% or 3.16 billion people around the globe would not be able to access healthcare facilities within one hour if they travel on foot. This figure drops to 8.9% or 646 million people with access to motorised transport.(29) Other studies in low- and middle-income countries (LMIC) settings have also shown that a significant share of individuals are not able to access healthcare in a timely manner.(30,31) Travel times are particularly higher in rural areas, with motorised transport availability and ownership improving access.(32–35) Similarly, in India, it has been observed that public healthcare accessibility is low in rural areas, particularly for those with no access to motorised transport. (36) However, studies on geographic access to public healthcare facilities in terms of travel time in the Indian context are few and limited in geographical breadth. The current study aims to estimate the travel times and the proportion of the population with timely geographic access to PHCs and CHCs in India.

## 2 Methods

### 2.1 Data sources

To comprehensively map the densities of PHCs and CHCs and their travel times, the study necessitated collecting, extracting, and collating data from multiple sources. The geolocation data for the PHCs and CHCs were extracted from the Geographic Information System (GIS) dataset of the Pradhan Mantri Gram Sadak Yojana (PMGSY).(37) The PMGSY is a wholly centrally sponsored scheme by the Government of India (GoI) to provide all-weather connectivity to unconnected habitations as a poverty alleviation strategy. We utilized the 2021 PMGSY GIS dataset and extracted geo-coordinates and other related geographical information for the health facilities under study, i.e., the PHCs and CHCs. PMGSY GIS data on PHCs and CHCs were not available for 7 states/UTs, i.e., Andaman & Nicobar Islands, Chandigarh, Dadra & Nagar Haveli and Daman & Diu (DNH and DD), Delhi, Lakshadweep, Puducherry, and Goa. At the district level, data on PHCs and CHCs were missing for 60 and 134 districts, respectively. The Malaria Atlas Project (MAP) data for the year 2019 were used to obtain the motorized and walking friction rasters for every square kilometre (1 km2).(38) GADM v3.6 data were used to delineate the administrative boundaries of India, while high-resolution (1 sq. km.) United Nations (UN) adjusted population counts from the WorldPop dataset for 2020 for India served as our source of population data.(39,40) The use of global data sources such as the MAP for travel time friction surfaces ensures that our findings are comparable to studies using these data sources from other countries and multi-country assessments. Validation of MAP-derived travel time rasters for certain geographies, including India, has been presented in original sources.(29,41)

### 2.2 Outcomes

The study had three outcomes. The first outcome was to calculate the density of health facilities, i.e., the number of PHCs and CHCs for every million population in India. The second outcome was to determine the median travel time to the closest PHC and CHC, from each 1 km2 raster in India. The third outcome was to estimate the proportion of the population with timely geographic access, also called Access Population Coverage (APC), to a PHC and a CHC. By walk, timely geographic access was defined as access within 30 minutes for PHCs and within 60 minutes for CHCs. For motorised transport, timely geographic access meant access within 60 minutes for PHCs, and 120 minutes for CHCs. These outcomes were calculated for India, 29 states and union territories (UTs), and 675 districts for PHCs and 601 districts for CHCs, and their urban and rural areas as well.

### 2.3. Data Analysis

For geocoding of the primary and secondary healthcare facilities (PHCs and CHCs), addresses were cleaned manually for improved machine readability. The Google Maps’ application programming interface (API) and the ‘Awesome Table’ add-on for Google Sheets for machine or API-based geocoding were used. For locations that returned multiple sets of coordinates, the ones with the most relevant address string were chosen. Geo-coordinates were used to identify and remove duplicates and points extending beyond the latitude and longitude limits of India.

An implementation of the Dijkstra algorithm was utilised to compute the minimum time required to traverse the friction surface from every 1 km^2^ pixel (grid cell) on the map to the geo-coordinates of every PHC and CHC. The algorithm was implemented for two modes of transport, i.e., walking and motorised transport. The proportion of the population with access to a PHC within 30 minutes by walking and 60 minutes by motorised transport, and the proportion with access to a CHC within 60 minutes by walking and within 120 minutes by motorised transport were the proxy variables for timely geographic access.

A binary accessibility raster was created with ‘1’s for pixels that fulfilled the timeliness criterion of each proxy variable and ‘0’s for cases otherwise. This raster was then overlaid on the population raster (extent matched). The population figures at each pixel were multiplied with the weights (i.e., 1s and 0s) to get the population number with timely geographic access. Proportions or percentages of the population with timely geographic access were obtained by dividing these numbers by the overall population.

Although PMGSY GIS data on PHCs and CHCs were missing for seven states/UTs and for a subset of 134 districts for CHC and 60 districts for PHC, these areas were retained during the computation of travel time and APC to preserve spatial continuity and consistency across geographies. However, to avoid reporting estimates based on incomplete facility data, all districts and states/UTs with missing PHC/CHC locations were explicitly masked in the outputs. Specifically, their values were set to NA, they were removed from all tabular summaries, and they were greyed out in all maps. As a result, no district- or state-level estimates are reported or interpreted for these areas, and they do not contribute to any reported summary statistics.

The workflow has been described in Figure 1. The same workflow has been discussed for investigating healthcare accessibility under the project, IndoHealMap.(42–47) Analysis was performed in R (Version 4.3.3).

**Figure 1:**
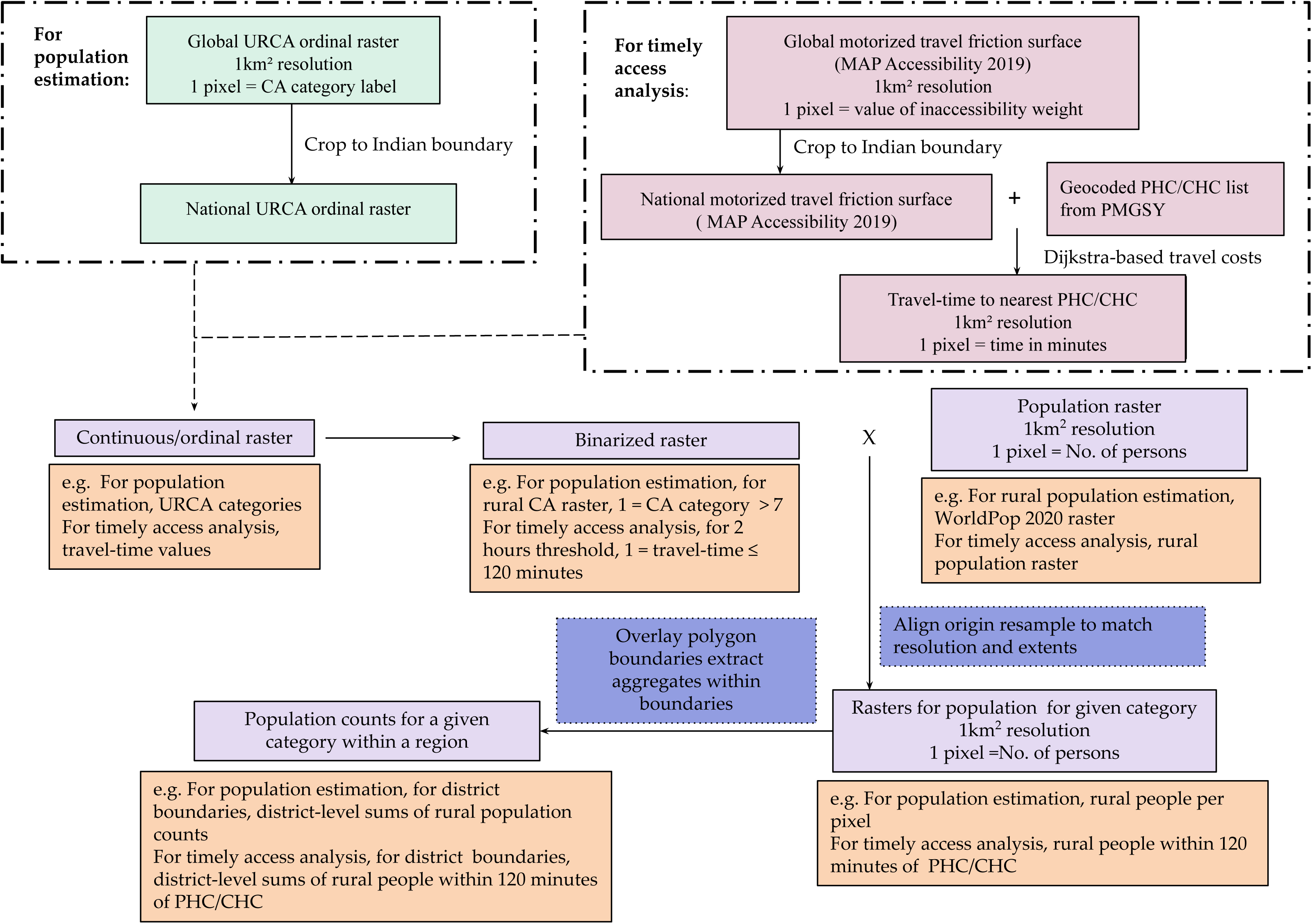
Raster-based estimation pipeline used for creating criteria-specific regional population counts.

## 3 Results

### 3.1 Density of healthcare facilities

#### 3.1.1 Primary Health Centres (PHCs)

According to the PMGSY GIS 2021 dataset, there were 31,958 PHCs in India with a national average density of 23.22 centres per million population (cpm). Among states/UTs, Maharashtra (3445) had the highest number of PHCs (3445) while the UT of Ladakh (31) had the lowest number of PHCs. The density of PHCs exceeded the national average in 25/29 (86.21%) states/UTs. The density of PHCs among states/UTs ranged from 8.46 cpm in West Bengal to 124.88 cpm in the state of Arunachal Pradesh. In rural areas, the density of PHCs ranged from 10.62 in West Bengal to 130.18 cpm in Arunachal Pradesh. In urban areas, the density ranged from 0 in 3 states (Arunachal Pradesh, Mizoram, and Ladakh) to 38.77 in Manipur. At the district level, the number of PHCs varied from 1 in Khagaria, Bihar, to 386 in Pune, Maharashtra. PHC density at the district level surpassed the national average in 391/675 (57.93%), and ranged from 0.49 cpm in Khagaria, Bihar, to 638.23 cpm in Anjaw, Arunachal Pradesh (**Figure 2**).

**Figure 2:**
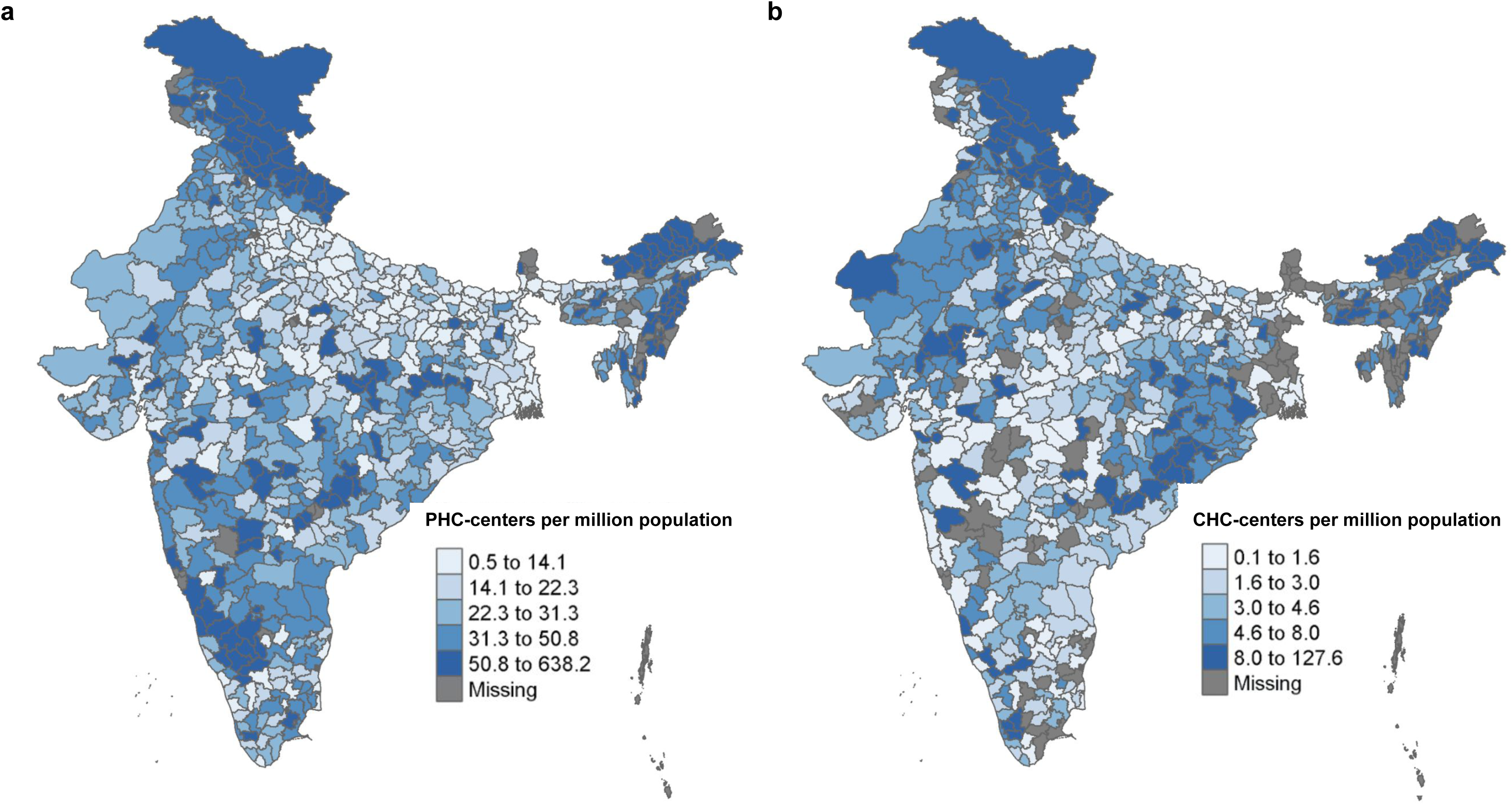
Map depicting the variation in density of a) PHCs and b) CHCs across districts in India.

#### 3.1.2 Community Health Centres (CHCs)

The total number of CHCs at the national level was 4237 centres. Among states/UTs, the number varied from 2 centres in Sikkim to 760 centres in Uttar Pradesh. The national average density of CHCs was estimated to be 3.07 cpm. The density of CHCs was found to be better than the national average in 16/29 (55.17%) states/UTs. The density of CHCs ranged from 0.16 cpm in West Bengal to 36.25 cpm in Arunachal Pradesh. In rural areas, the density ranged from 0.13 cpm in West Bengal to 37.2 in Arunachal Pradesh. In urban areas, the density ranged from 0 in Ladakh to 14.3 in Arunachal Pradesh. At the district level, the number of CHCs varied from 1 in 71/601 (11.83%) districts to 95 in Ahmednagar, Maharashtra. The density at the district level exceeded the national average in 353/601 (58.74%). The lowest and the highest density of PHCs were 0.08 cpm in South Twenty Four Parganas and 127.64 cpm in Anjaw, Arunachal Pradesh **(Figure 2)**.

### 3.2 Travel time to the nearest PHC

#### 3.2.1 By walking

At the national level, the median time taken to access a PHC by walking was found to be 79.53 (IQR: 45.88-134.00) minutes. The travel time was shorter than the national median in 15/29 (51.72%) states/UTs. The travel times ranged from 47.95 (IQR: 28.86-73.05) minutes in Punjab to 852.32 (IQR: 477.10-1338.35) minutes in the UT of Ladakh. At the district level, 409/675 (60.59%) districts had a shorter travel time by walking compared to the national value. The walking time to a PHC was the shortest in Nalanda, Bihar [25.06 (IQR: 14.97-41.02) minutes] and the longest in Leh, Ladakh [858.47 (IQR: 501.86-1359.10) minutes]. The travel times for rural and urban regions are summarized in **Table 1**.

**Table 1.** Summary of travel times for rural and urban regions.

| **National level** |  |  |  |  |
| --- | --- | --- | --- | --- |
| Median (IQR) travel time (in minutes) |  |  |  |  |
|  | PHC (Rural) | PHC (Urban) | CHC (Rural) | CHC (Urban) |
| Walking | 79.8 (46.12-134.28) | 60.77 (34.66-94.58) | 228.36 (131.55-384.50) | 173.65 (96.94-275.93) |
| Motorised transport | 17.16 (8.04-35.61) | 6.99 (3.82-11.22) | 36.80 (20.68-64.36) | 18.26 (10.55-29.29) |
| **State/UT level** |  |  |  |  |
| States/UTs with travel time shorter than national median, n (%) |  |  |  |  |
|  | PHC (Rural) | PHC (Urban) | CHC (Rural) | CHC (Urban) |
| Walking | 14/29 (48.28%) | 18/29 (62.07%) | 15/29 (51.72%) | 15/29 (51.72%) |
| Motorised transport | 12/29 (41.38%) | 15/29 (51.72%) | 14/29 (48.28%) | 15/29 (51.72%) |
| Shortest median (IQR) travel time (in minutes) |  |  |  |  |

|  | PHC (Rural) | PHC (Urban) | CHC (Rural) | CHC (Urban) |
| --- | --- | --- | --- | --- |
| Walking | Punjab:<br>47.92 (28.76 - 72.84) | Manipur:<br>25.11 (11.08 - 35.24) | Kerala:<br>112.31 (64.29-199.52) | Kerala:<br>88.92 (57.80-128.72) |
| Motorised transport | Kerala:<br>6.97 (3.79-17.91) | Manipur:<br>2.50 (1.58 - 4.14) | Kerala:<br>13.69 (7.77-30.88) | Kerala:<br>9.39 (6.15-13.48) |
| Longest median (IQR) travel time (in minutes) |  |  |  |  |
|  | PHC (Rural) | PHC (Urban) | CHC (Rural) | CHC (Urban) |
| Walking | Ladakh:<br>852.06 (477.00 -1337.78) | Mizoram:<br>154.12 (133.73-170.86) | Ladakh:<br>1773.88 (1027.79-2513.11) | Mizoram:<br>863.19 (450.79-897.74) |
| Motorised transport | Ladakh:<br>311.74 (126.63-601.20) | Mizoram:<br>11.70 (9.83-13.55) | Ladakh:<br>396.73 (218.88-681.95) | Mizoram:<br>65.78 (40.40-27.88) |
| **District level** |  |  |  |  |
| Districts with travel time shorter than the national median, n (%) |  |  |  |  |
| Walking | 410/674 (60.83%) | 261/504 (51.78%) | 390/565 (69.02%) | 250/387 (64.60%) |
| Motorised transport | 406/674 (60.24%) | 281/504 (55.75%) | 376/565 (66.55%) | 257/387 (66.41%) |
| Shortest median (IQR) travel time (in minutes) |  |  |  |  |
|  | PHC (Rural) | PHC (Urban) | CHC (Rural) | CHC (Urban) |
| Walking | Nalanda, Bihar:<br>25.06 (14.97-40.30) | Bishnupur, Manipur:<br>11.07 (10.12-20.23) | Alappuzha, Kerala:<br>57.74 (37.36-84.46) | Changlang, ARP<sup>2</sup>:<br>13.60 (9.88-21.03) |
| Motorised transport | Pulwama, J&K<sup>1</sup>:<br>2.87 (1.56-32.54) | Bishnupur, Manipur:<br>1.63 (0.79-2.41) | Alappuzha, Kerala:<br>6.7 (4.11-10.01) | Tuensang, Nagaland:<br>1.68 (0.86-1.24) |
| Longest median (IQR) travel time (in minutes) |  |  |  |  |
|  | PHC (Rural) | PHC (Urban) | CHC (Rural) | CHC (Urban) |
| Walking | Leh, Ladakh:<br>858.06 (501.72-1358.29) | Singrauli, MP<sup>3</sup>:<br>176.79 (130.21-196.24) | Leh, Ladakh:<br>2059.70 (1487.61-2706.89) | Dhule, Maharashtra:<br>862.40 (422.29-888.83) |
| Motorised transport | Anjaw, ARP<sup>2</sup>:<br>600.27 (316.82-965.44) | Punch, J&K<sup>1</sup>:<br>22.12 (2.62-68.03) | KK<sup>4</sup>, ARP<sup>2</sup>:<br>1007.826 (437.17-1558.69) | Dhule, Maharashtra<br>67.61 (45.15-70.43) |

<sup>1</sup>J&K: Jammu & Kashmir
<sup>2</sup>ARP: Arunachal Pradesh
<sup>3</sup>MP: Madhya Pradesh
<sup>4</sup>KK: Kurung Kumey

#### 3.2.2 By motorised transport

At the national level, the median time taken to access a PHC by motorized transport was found to be 16.77 (IQR: 7.79-35.10) minutes. Among states/UTs, 13/29 (44.82%) had a shorter travel time compared to the national median. The state of Kerala had the shortest median travel time, 6.39 (IQR: 3.56-13.64) minutes, and the UT of Ladakh had the longest time, 311.80 (IQR: 126.65-601.51) minutes. At the district level, the travel time was shorter than the national median in 413/675 (61.18%) districts. The travel time ranged from 2.68 (IQR: 1.48-28.98) minutes in Pulwama, Jammu & Kashmir, to 604.11 (IQR: 318.51-650.93) minutes in Anjaw, Arunachal Pradesh. The travel times for rural and urban regions are summarized in **Table 1**.

### 3.3 Median travel time to the nearest CHC

#### 3.3.1 By walking

At the national level, the median time to reach the nearest CHC by walk was found to be 227.53 (IQR: 130.80-384.07) minutes. At the state/UT level, 14/29 (48.27%) had a shorter travel time compared to the national median. The median travel time varied from 107.40 (IQR: 64.04-183.95) minutes in Kerala to 1774 (IQR: 1028.23-2513.59) minutes in the UT of Ladakh. The median travel time was shorter than the national value in 399/601 (66.39%) districts. The median time on foot was found to be the shortest in the Bongaigaon district of Assam at 61.63 (IQR: 37.22-92.41) minutes and the longest in the Leh district [2059.95 (IQR: 1488.33-2707.40) minutes] of Ladakh. The travel times for rural and urban regions are summarized in **Table 1**.

#### 3.3.2 By motorised transport

At the national level, the median time to access a CHC by motorised transport was estimated to be 36.23 (IQR: 20.22-63.92) minutes. The travel time was shorter than the national median in 14/29 (48.27%) states/UTs. The median time ranged from 12.66 (IQR: 7.44-25.28) minutes in Kerala to 396.83 (IQR: 218.94-682.48) minutes in Ladakh. The median travel time was shorter than the national value in 395/601 (65.72%) districts. The median time in districts varied from 7.58 (IQR: 5.03-10.49) minutes in Kottayam, Kerala, to 1008.83 (IQR: 437.32-1560.25) minutes in Kurung Kumey of Arunachal Pradesh. The travel times for rural and urban regions are summarized in **Table 1**.

### 3.4 Access Population Coverage (APC) for PHCs

#### 3.4.1 By walking

By walking, timely geographic access to a PHC meant being able to access one within 30 minutes. At the national level, the APC was estimated to be 18.69%. At the state/UT level, 14/29 (48.28%) had an APC greater than the national median. APC ranged from 5.2% in Mizoram to 43.71% in Manipur. Among districts, 297/675 (44%) had an APC greater than the national average, and greater than 50% in 18/675 (2.66%) districts. APC varied from 0.78% in Upper Siang, Arunachal Pradesh, to 69.68% in Bishnupur, Manipur **(Figure 3)**. APC was less than 50% in all states/UTs. The APCs for urban and rural regions have been summarized in **Table 2**. APC was less than 50% in all rural and urban areas except urban Manipur (69.3%). APC was 0% in urban Mizoram.

**Figure 3:**
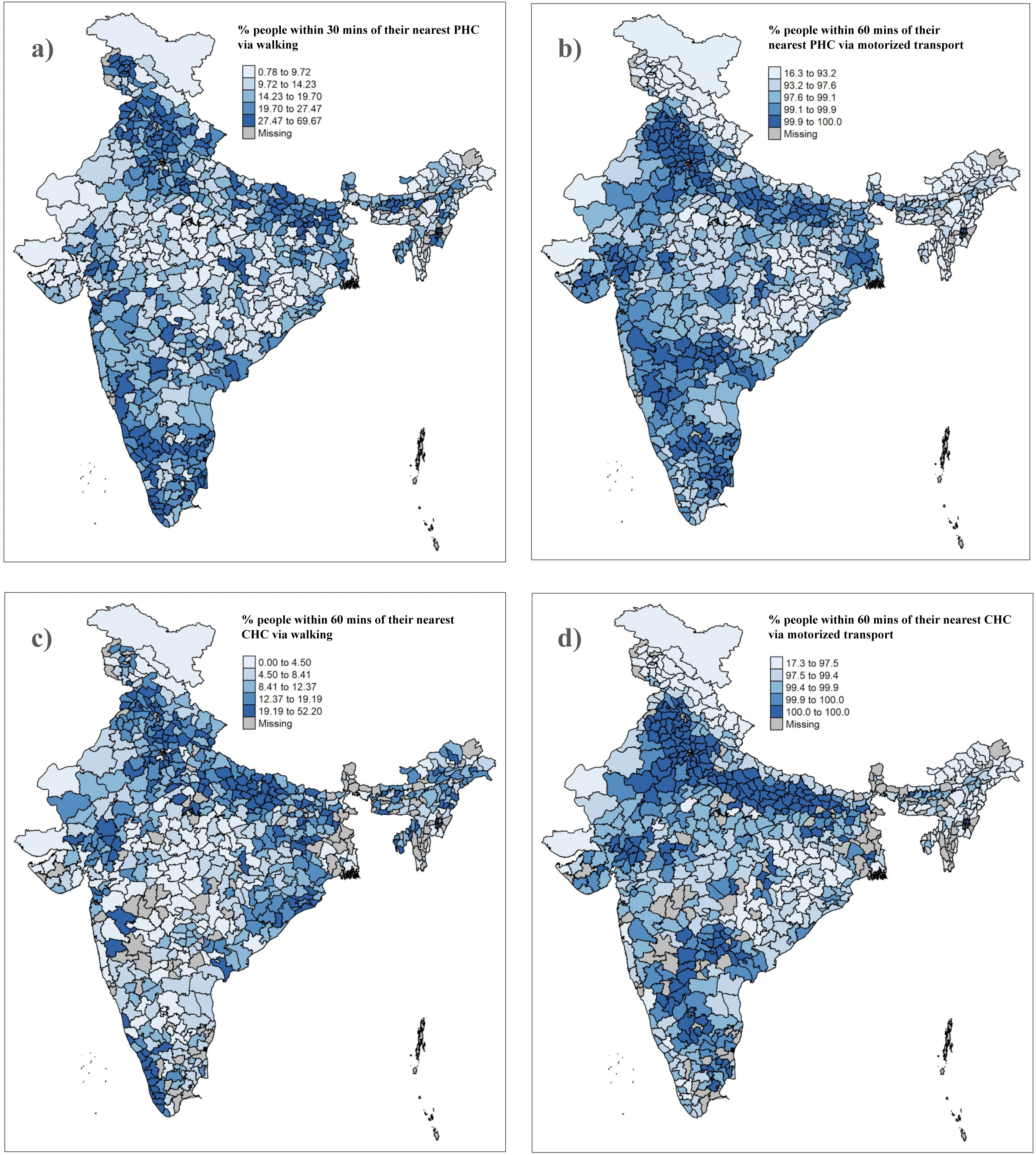
Maps depicting the APC in the districts of India for a) PHCs by walking, b) PHCs by motorised transport, c) CHCs by walking, and d) CHCs by motorised transport.

**Table 2.**
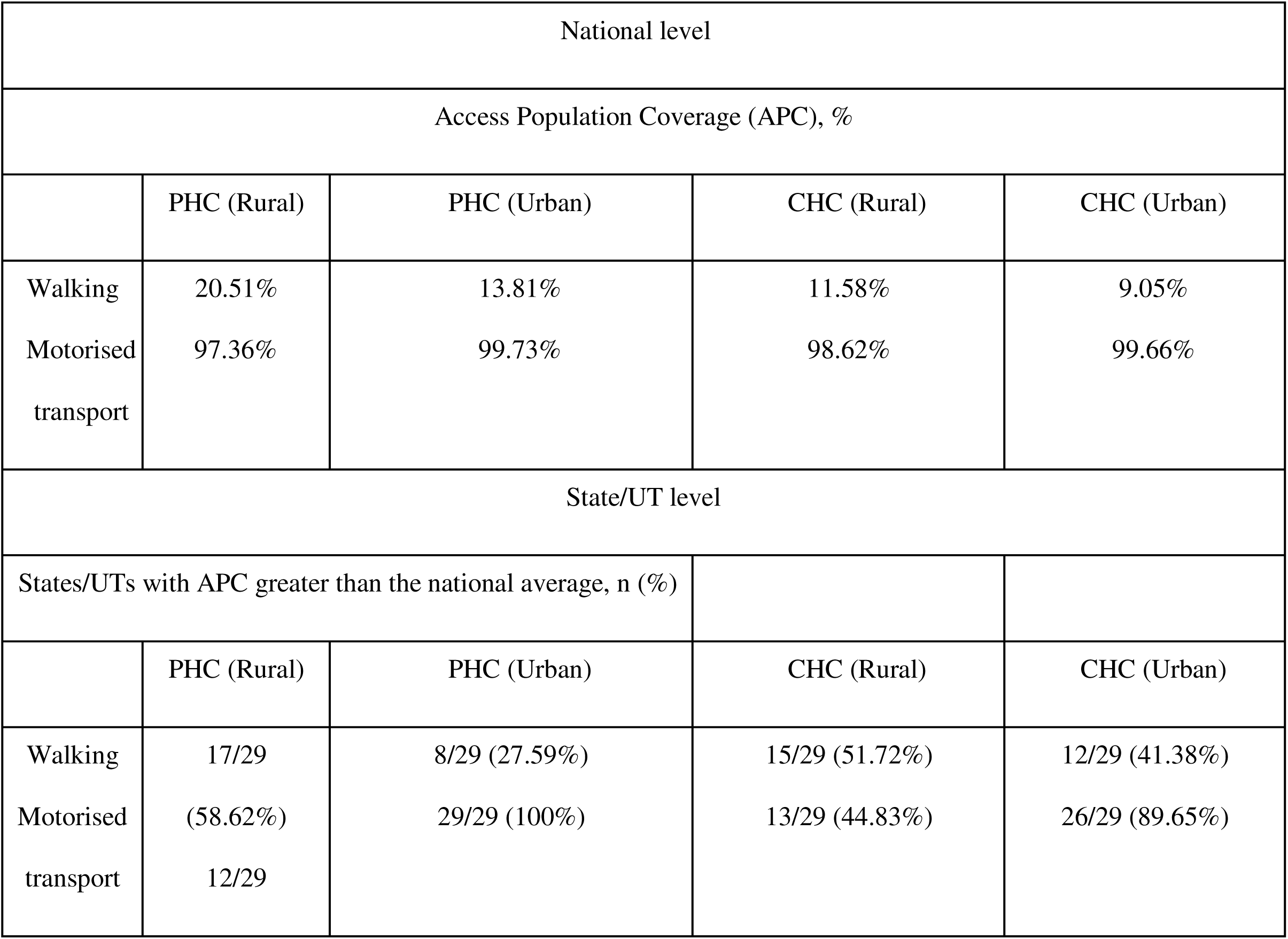
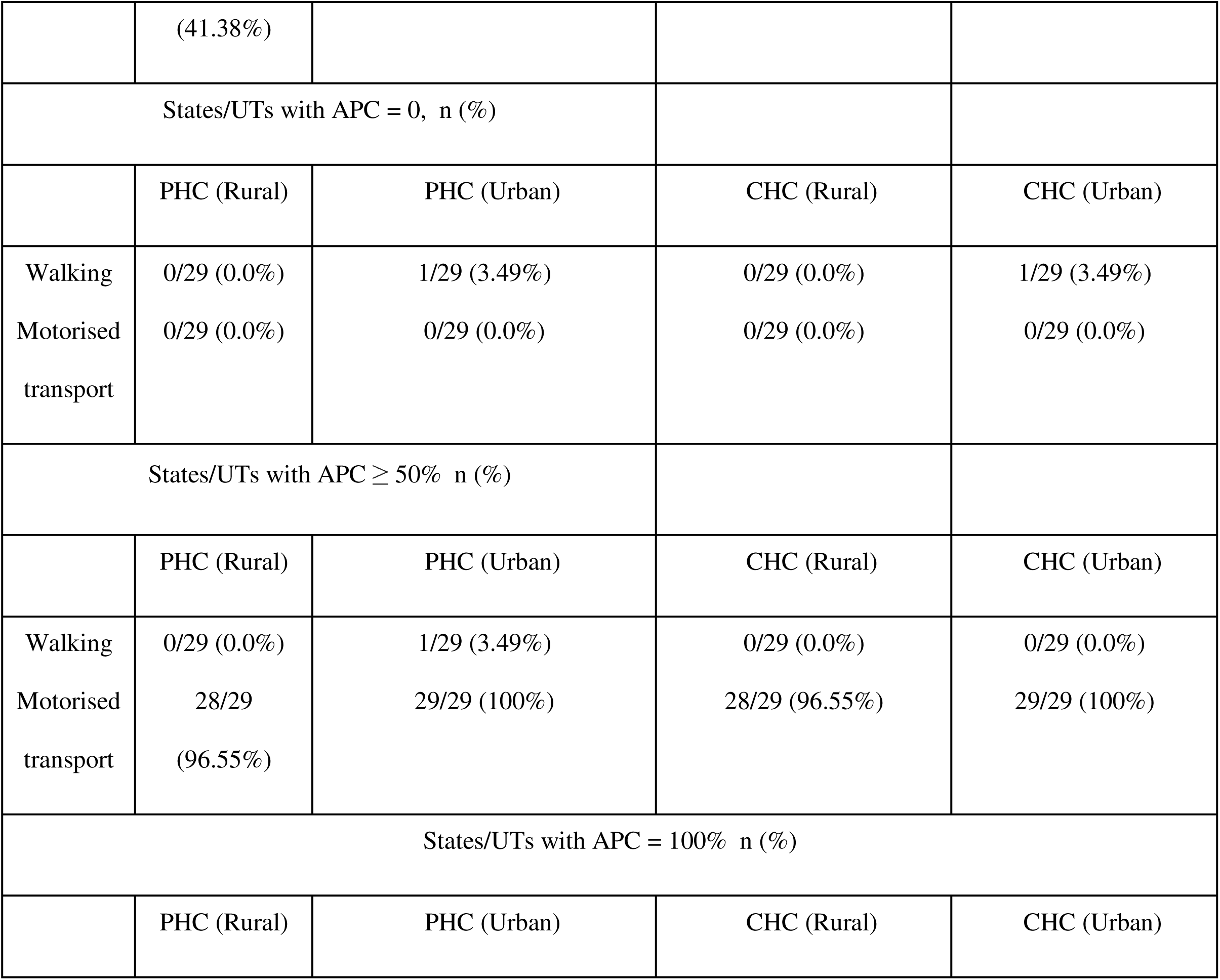
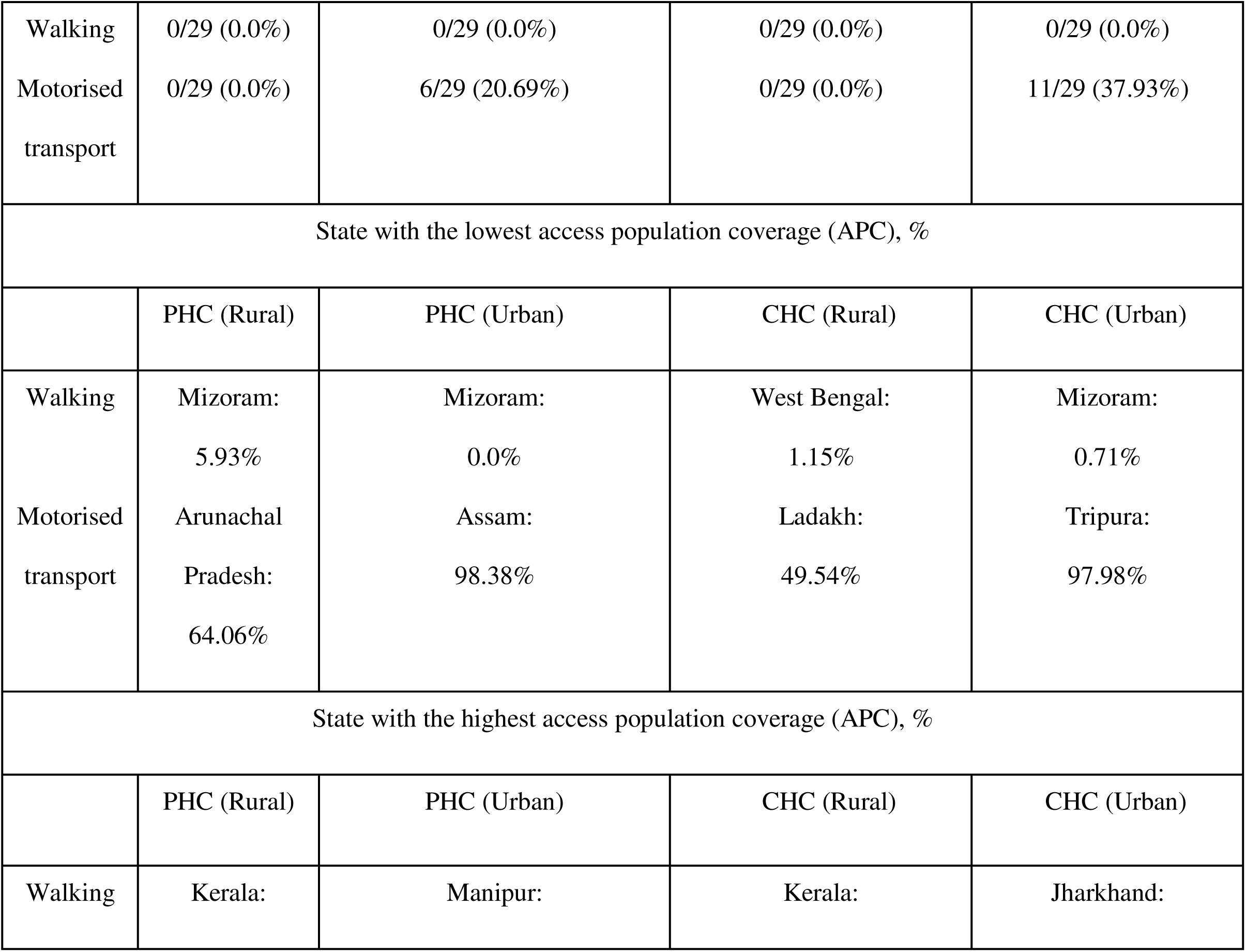
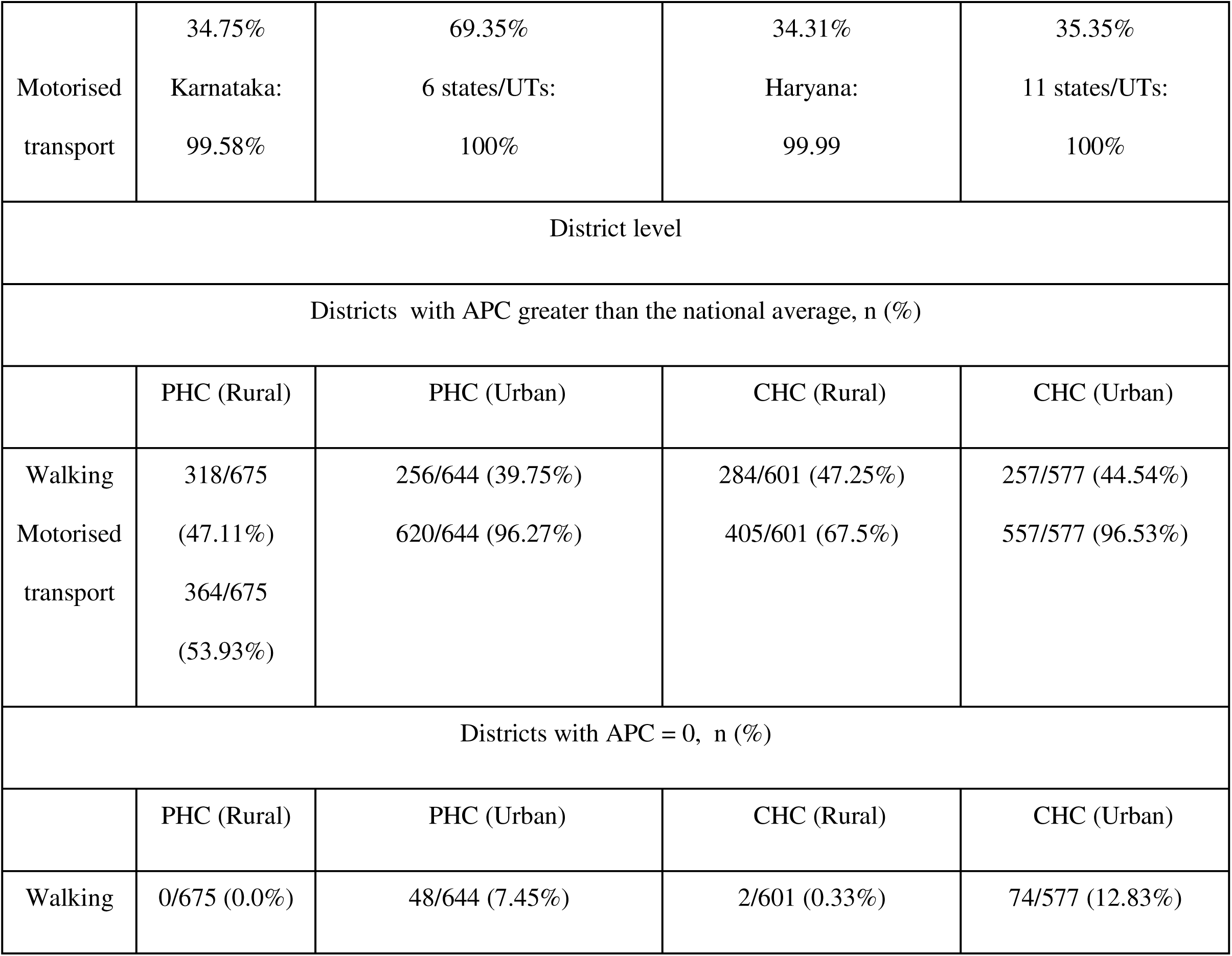
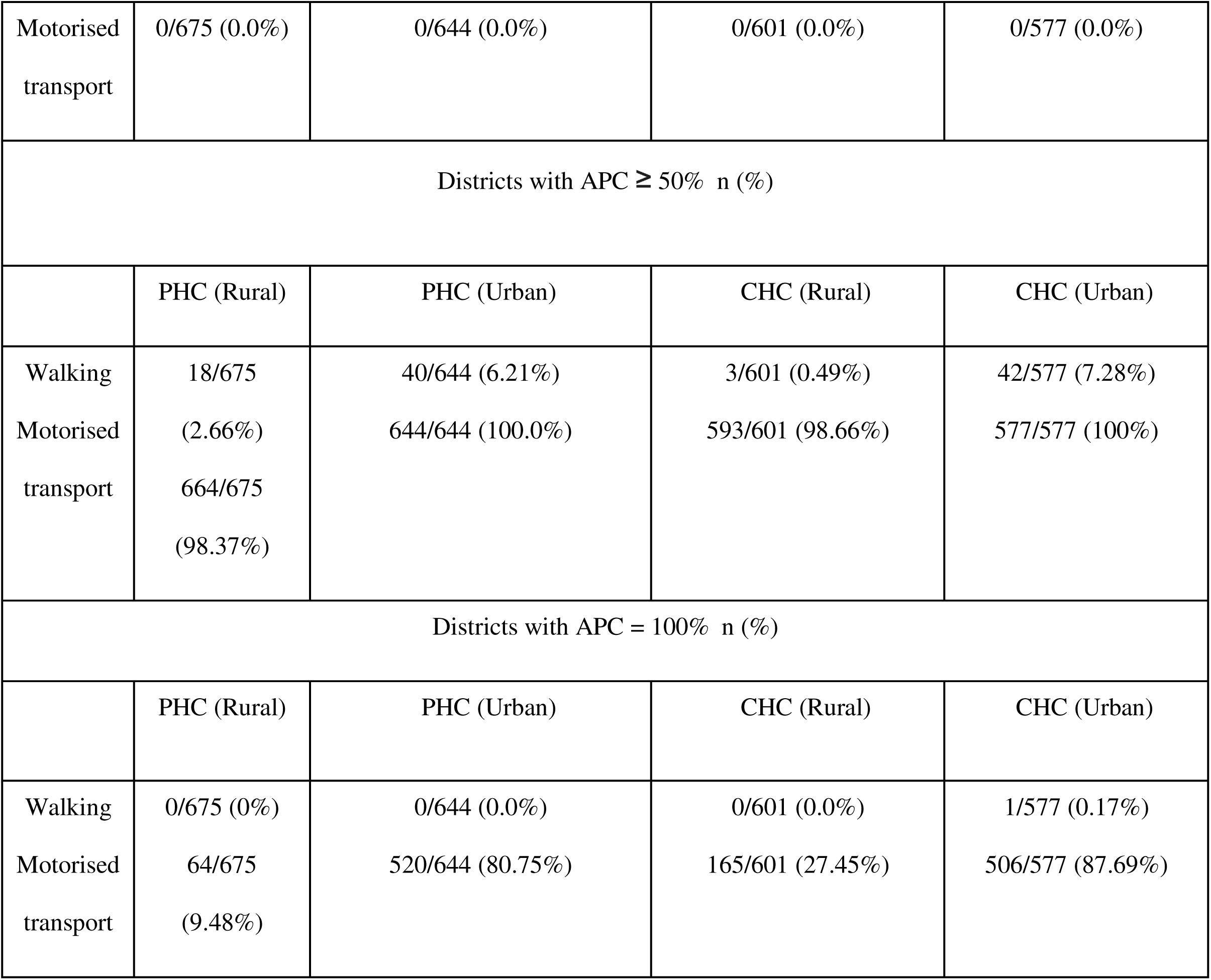
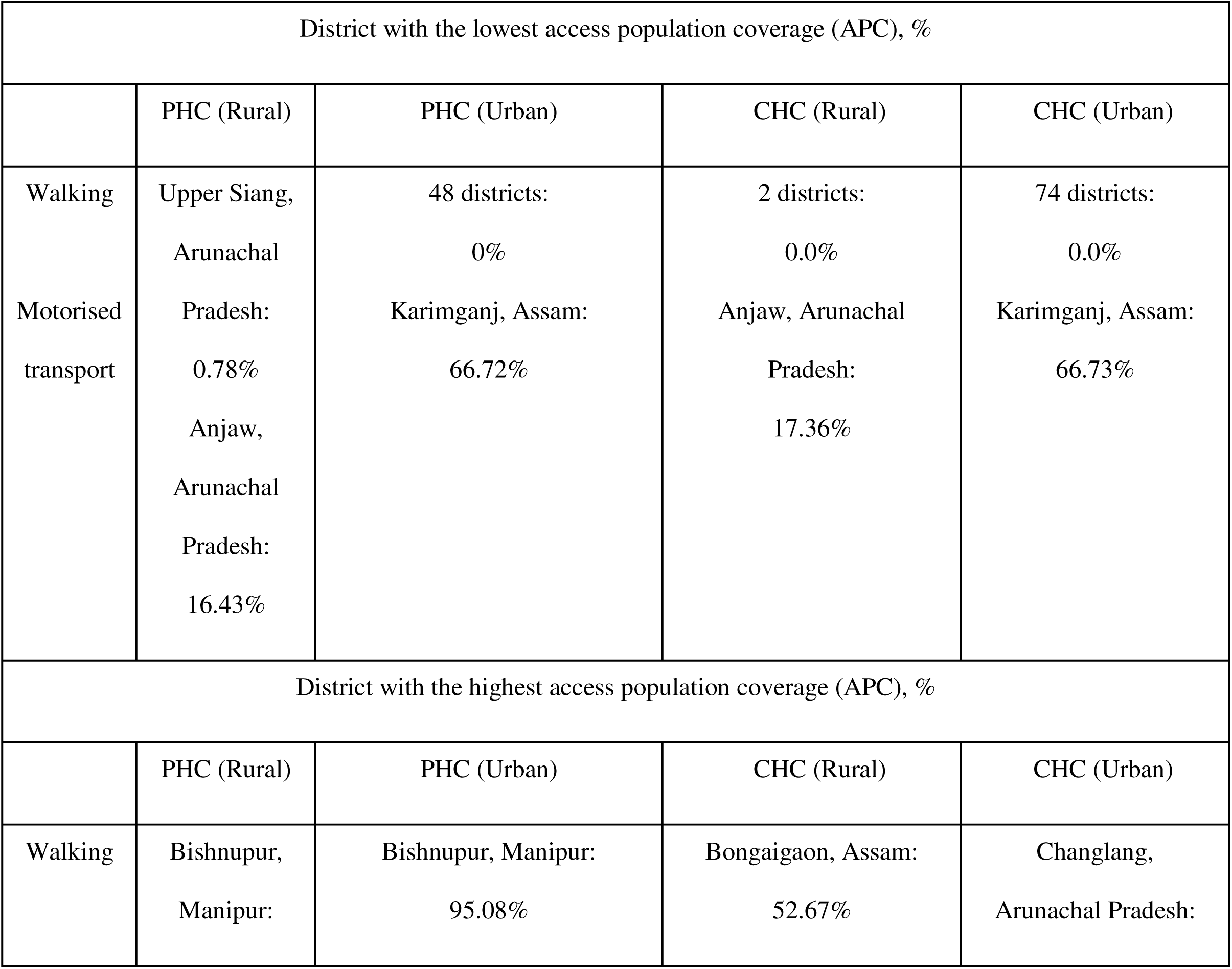
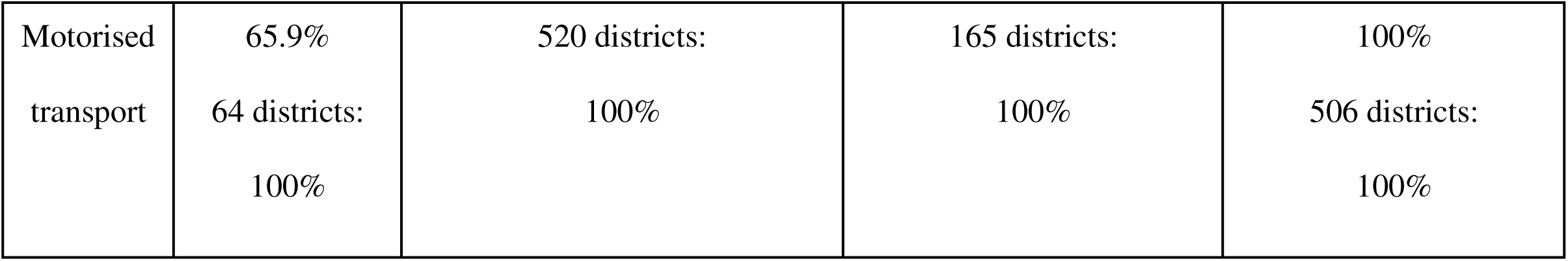
Summary of access population coverage (APC) for rural and urban regions.

#### 3.4.2 By motorised transport

By motorised transport, timely geographic access to a PHC meant being able to access one within 60 minutes. At the national level, the APC was estimated to be 97.97%. At the state/UT level, 13/29 (44.83%) states/UTs had an APC greater than the national average. APC was greater than 50% in 28/29 (96.55%) states/UTs, and ranged from 44.70% in Ladakh to 99.98% in Haryana. APC was greater than the national average in 382/675 (56.59%) districts and greater than 50% in 664/675 (98.37%) districts. APC ranged from 13.39% in Anjaw, Arunachal Pradesh, to 100% in 64/675 (9.48%) districts **(Figure 3)**. The APCs for urban and rural regions have been summarized in **Table 2**. APC was greater than 50% in urban areas of all states/UTs. In rural areas, the APC was less than 50% in Ladakh (44.6%).

### 3.5. Access Population Coverage (APC) for CHCs

#### 3.5.1 By walking

By walking, timely geographic access to a CHC meant being able to access one within 60 minutes. At the national level, the APC was estimated to be 10.89%. At the state/UT level, 13/29 (44.83%) had an APC greater than the national average. APC ranged from 1.5% in West Bengal to 29.39% for the state of Kerala. Among districts, 277/601 (46.09%) had an APC greater than the national average, while APC was found to be greater than 50% in 2/601 (0.33%) districts. APC varied from 0% in 2/601 (0.33%) districts to 52.20% in Dhanbad, Jharkhand **(Figure 3)**. APC was estimated to be 57/731 (7.8%) districts. The APCs for urban and rural regions have been summarized in **Table 2**. APC was less than 1% in urban Mizoram.

#### 3.5.2 By motorised transport

By motorised transport, timely geographic access to a CHC meant being able to access one within 120 minutes. At the national level, the APC was estimated to be 98.88%. Among states/UTs, APC was greater than the national average in 13/29 (44.83%) states/UTs and greater than 50% in 28/29 (96.55%) states/UTs, respectively, and ranged from 49.65% in Ladakh to 99.99 in Haryana. Among districts, 416/601 (69.22%) had an APC greater than the national average, while APC was found to be greater than 50% in 593/601 (98.67%) districts. The APCs for urban and rural regions have been summarized in **Table 2**.

## 4 Discussion

### 4.1 Summary and interpretation of findings

The main aim of this study was to map timely geographic access to primary and secondary public healthcare facilities across geographic scales from the national to the district levels in India. Arunachal Pradesh, Himachal Pradesh, Nagaland, and Ladakh were observed to have the highest densities of PHCs and CHCs. The lowest densities for PHCs were seen in West Bengal, Bihar, and Uttar Pradesh, and the lowest densities for CHCs were observed in West Bengal, Tamil Nadu, and Maharashtra. Travel times by walking were the longest in states/UTs with difficult terrain, including Arunachal Pradesh, Ladakh, Mizoram, and Sikkim. Given that rural areas have greater PHCs and CHCs per million than urban areas, urban areas fared better than rural areas for median travel time, possibly due to a dearth of pedestrian-friendly infrastructure in rural areas. The average road density for India in 2018-19 was 5296.3 per 1000 km^2^ for urban areas in comparison to 1458.1 for rural areas.(48) This could explain the relatively shorter travel times observed for urban areas by motorised transport.

However, even in the case of motorised transport, travel times were the longest in the states/UTs of Arunachal Pradesh, Ladakh, Mizoram, and Sikkim. Therefore, it can be seen that the nature of the terrain in these regions tends to slow down movement in general, whether on foot or by motorised transport. Travel time, a summary statistic, cannot showcase a holistic picture of geographic access. Hence, we looked at access population coverage (APC) as a measure of geographic access. Geographic access by walking was found to be poor. Overall, approximately 1118 million or 81.31% Indians, 798 million or 79.49% rural Indians, and 320 million or 86.19% urban Indians were beyond the 30-minute threshold. Similarly, approximately 1225 million or 89.11% Indians, 888 million or 88.42% rural Indians, and 337 million or 90.95% urban Indians were beyond the 60-minute threshold for CHCs. Geographic access by walking was low in Arunachal Pradesh, Ladakh, and Mizoram. This could have resulted from the long travel times in Arunachal Pradesh, Ladakh, and Mizoram. Geographic access by walking was heterogeneous across states/UTs and districts, but was generally better in rural areas compared to urban areas, despite the longer travel times. This could be credited to improvements in the rural health infrastructure brought about by sustained investments over the last two decades through the National Rural Health Mission (NRHM).(49) On the whole, these figures depict an alarming situation, as they indicate that hundreds of millions of Indians lack geographic access to primary and secondary healthcare, resulting in significant repercussions for their health.

Approximately 28 million Indians, 27 million rural Indians, and 1 million urban Indians were still beyond the 60-minute threshold by motorised transport for PHCs. For CHCs, approximately 15 million, 14 million rural Indians, and 1 million urban Indians were beyond the 120-minute threshold. Although these percentages appear small, they translate into a sizable number of Indians lacking geographic access and merit a deeper look for action. Geographic access was on the lower end for Ladakh, Arunachal Pradesh, and Mizoram, possibly due to the long travel times in these regions. The small administrative areas of these UTs and well-developed road networks, due to their urban nature, could have facilitated quick access to primary and secondary healthcare facilities beyond their borders. There was marked heterogeneity in geographic access by motorised transport across states/UTs and districts, but it was generally better in urban areas.

### 4.2 Strengths and limitations

Our study has the following strengths. First, using the PMGSY GIS dataset allowed us to analyse precisely geocoded locations of primary and secondary healthcare facilities. This enabled us to comprehensively map travel times and geographic access using APC. Second, travel times and APCs were estimated for two modes of travel: walking and motorised transport, and two regions: rural and urban. Third, estimates were calculated up to the district level, lending further strength to our study.

There were certain limitations as well. First, data related to primary and secondary health facilities were not available for several states/UTs and districts. Second, our study is also limited by the lack of information on the functional status of the health facilities. Third, the estimates for walking do not account for groups like the elderly or disabled, who would lack access despite geographical and temporal proximity. Fourth, the APCs for motorised transport assume that everybody has access to it, the unavailability of which would preclude an individual from accessing a health facility on time. Fifth, the travel time raster data from MAP is from 2019. India has observed remarkable growth in roadway infrastructure, which can potentially reduce travel times and improve APC. Hence, our estimates for travel time estimates are likely higher, and those for APC values are lower than India’s current reality. In other words, we present a conservative picture of timely geographic access. However, we used the most recent available travel-time raster data and updated facility data. Future studies can use similar methods to update the estimates, once travel time rasters are available for 2025. Sixth, since we did not have access to resource metrics required to inform model weights, such as the number of beds, healthcare workers, or surgical volumes, we used a parsimonious approach instead of advanced models like Floating Catchment Area (FCA) or Rational Agent Modeling (RAM). Future studies should employ FCA or RAM to assess measures of geographic access that include resource metrics. Seventh, due to limited data on time thresholds, we adopted arbitrary thresholds. Most literature in the past has focused on a distance-based threshold. The Lancet Commission on Global Surgery suggested a threshold of 120 minutes was associated with reduced maternal mortality.(50) This guided our selection of the motorized travel time threshold for CHCs, where institutional deliveries are a key service. The rationale for the remaining thresholds was two-fold: first, they must be less than 120 minutes; and second, the access to PHCs should be higher than that to CHCs, as the former are primary-level care facilities. Lastly, our approach used either walking or motorized transport. We acknowledge that travel to a healthcare facility often necessitates more complex of different transportation modes. However, travel time raster data to consider such complex travel scenarios remains unavailable. Hence, our analyses may not reflect the realistic travel patterns in India. Even so, the walking and motorized modes of transport provide the upper and lower bounds of travel times. Travel time could be as short as that of motorized transport, assuming optimal conditions, or as long as the time required to walk to the nearest healthcare facility. The true travel time would lie between the two bounds. Contingent on data availability, future studies should explore hybrid travel models for a more accurate assessment of timely geographic access.

### 4.3 Implications and Takeaways

Increased travel times and low geographic access can result in reduced utilization of health services and worsen health outcomes. Issues with healthcare access also impede the pursuit of Universal Health Coverage (UHC), and deepen inequities.(9,13–15,18–22,24,30) We identified three states/UTs that must be the focus of future policy and efforts like APC-guided facility placement and improvements in pedestrian and road infrastructure, as they were consistently found to have long travel times and low geographic access: Arunachal Pradesh, Mizoram, and Ladakh. Our findings indicate that the current spatial distribution of primary and secondary healthcare facilities in India does not favour geographic access for a major portion of the Indian population by walking. A spatial coverage modelling study done in one district of Jammu & Kashmir, India, reported that access to ambulatory and immunization care within one hour of walking was 17%.(36) A similar study from Western Province, Rwanda, found that 26.6% of the population could access the primary health care network by walking within one hour.(34) This divergence in findings across studies can be explained by the differences in study settings and the methodologies used to determine geographic access. Adding new healthcare facilities or strategically relocating existing ones, using APC as a guidance tool, can improve walkability.

Geographic access by motorised transport to primary and secondary healthcare facilities in the current study was high. This phenomenon of high geographic access by motorised transport has been noted in other studies as well.(29,33,36) However, geographic access was lower in rural areas compared to urban areas. This can be addressed by improving public road transport and strengthening ambulance networks in these areas. These measures are faster to implement and are a pragmatic stopgap until a more equitable APC-guided establishment of future healthcare facilities can be achieved over a long-term horizon. While improving timely geographic access through enhancements to public road transport and ambulance networks is a crucial, faster-to-implement measure, it primarily addresses the resistance in transport, not the fundamental lack of service availability. For timely geographic access, the proximity and availability of facilities remain the most decisive factors in increasing utilization and achieving true timely geographic access. Therefore, the long-term, structural solution remains the establishment of new facilities or optimization of locations guided by APC. Furthermore, our study also lends support to the use of APC as a measure of geographic access over travel times. We noticed that the travel times for walking in urban areas were shorter compared to rural areas, but APC values were lower for urban areas. This is because APC takes into account population size, density, and spatial distribution of health facilities, providing a more holistic view of geographic access. Finally, it is necessary to highlight that future research must incorporate the functional status of the health facilities as well as population attributes that facilitate or impede geographic access. This will result in more precise estimates of geographic access to health facilities and better inform policy making and programmes.

## 5 Conclusion

This study finds several gaps in timely geographic access to primary and secondary healthcare facilities in India, particularly for walking. Arunachal Pradesh, Mizoram, and Ladakh, with long travel times and low geographic access, should be the priority areas for future interventions. Focusing on APC-based facility establishment and pedestrian infrastructure can help improve timely geographic access by walking. Additionally, rural areas must be prioritized for improvements in road transport and ambulance networks. Addressing these disparities in timely geographic access is essential for improving health outcomes and achieving UHC.

## Data Availability

All data produced in the present study are available upon reasonable request to the authors

## 6 List of Abbreviations

PHC: Primary Health Centre
CHC: Community Health Centre
IPHS: Indian Public Health Standards
LMIC: Low-and Middle-Income Countries
GIS: Geographic Information System
PMGSY: Pradhan Mantri Gram Sadak Yojana
GoI: Government of India
MAP: Malaria Atlas Project
UN: United Nations
APC: Access Population Coverage
UT: Union Territory
API: Application Programming Interface
RHS: Rural Health Statistics

## 7 Declarations

### Ethics approval and consent to participate

Not applicable

### Consent for publication

Not applicable

### Availability of data and materials

The datasets used and/or analysed during the current

study is available from the corresponding author on reasonable request.

## Authors’ contributions

### Study concept and design

Siddhesh Zadey

### Acquisition, analysis, or interpretation of data

Shreyas Patil, Harsh Thakkar, Aamir Miyajiwala, Anoushka Arora, Harsh Thakkar, Ankit Raj, Siddhesh Zadey

### Drafting of the manuscript

Shreyas Patil, Anoushka Arora

### Critical revision of the manuscript for important intellectual content

All authors

### Statistical analysis

Aamir Miyajiwala, Harsh Thakkar, Ankit Raj, Siddhesh Zadey

### Administrative, technical, or material support

Siddhesh Zadey

### Study supervision

Siddhesh Zadey

## Acknowledgements

None

